# Evaluating Loon Lens Pro™, an AI-Driven Tool for Full-Text Screening in Systematic Reviews: A Validation Study

**DOI:** 10.1101/2025.02.11.25322087

**Authors:** Ghayath Janoudi, Mara Uzun (Rada), Mia Jurdana, Brian Hutton

## Abstract

**Background:** Systematic literature reviews (SLRs) are essential for evidence synthesis but are hampered by the resource-intensive full-text screening phase. Loon Lens Pro™, a publicly available agentic AI tool, automates full-text screening without prior training by using user-defined inclusion/exclusion criteria and multiple specialized AI agents. This study validated Loon Lens Pro™ against human reviewers to assess its accuracy, efficiency, and confidence scoring in screening.

**Methods:** In this comparative validation study, 84 full-text articles from eight SLRs were screened by both Loon Lens Pro™ and human reviewers (gold standard). The AI provided binary inclusion/exclusion decisions along with a transparent rationale and confidence ratings (low, medium, high). Performance metrics— including accuracy, sensitivity, specificity, negative predictive value, precision, and F1 score—were derived from a confusion matrix. Logistic regression with bootstrap resampling (1,000 iterations) evaluated the association between confidence scores and screening errors.

**Results:** Loon Lens Pro™ correctly classified 70 of 84 full texts, achieving an accuracy of 83.3% (95% CI: 75.0– 90.5%), sensitivity of 94.7% (95% CI: 82.4–100%), and specificity of 80.0% (95% CI: 70.1–89.2%). The negative predictive value was 98.1% (95% CI: 93.8–100%), with a precision of 58.1% (95% CI: 41.4– 76.0%) and an F1 score of 0.72. Logistic regression revealed a strong inverse relationship between confidence level and error probability: low, medium, and high confidence decisions were associated with predicted error probabilities of 46.9%, 30.9%, and 3.5%, respectively (C-index = 0.87).

**Conclusion:** Our study provides evidence that Loon Lens Pro™ is a viable and effective tool for automating the full-text screening phase of systematic reviews. Its high sensitivity, robust confidence scoring mechanism, and transparent rationale generation collectively support its potential to alleviate the burden of manual screening without compromising the quality of study selection.

## Background

Systematic literature reviews (SLRs) play a pivotal role in synthesizing evidence across disciplines, offering comprehensive insights that inform clinical practice, guideline development, and policymaking. SLRs are especially critical in fields experiencing rapid growth in research output, where the accumulation of evidence demands rigorous and transparent methodologies to ensure reliability. However, the SLR process, particularly the full-text screening stage, remains resource-intensive, requiring considerable time, labor, and expertise.^1–3^ It is estimated that the screening phase of systematic reviews consumes about 25% of the total effort per review.^4^

The full-text screening phase, also known as level 2 (L2) screening, involves evaluating potentially eligible full text articles against predefined inclusion and exclusion criteria. This process can entail the detailed review of hundreds of full-text documents, contributing significantly to the overall workload and time required to complete a systematic review. Challenges such as reviewer fatigue, inconsistencies in decision-making, and the sheer volume of literature exacerbate the difficulty of maintaining accuracy and efficiency in the process of study selection.^2^ Given the importance of complete study identification in SLRs of comparative efficacy and safety, methods to address these challenges have long been of interest to the knowledge synthesis community.

Recent advances in artificial intelligence (AI) and large language models (LLMs) have introduced promising solutions to enhance the efficiency of performing systematic reviews. AI-driven tools have demonstrated potential in earlier stages of systematic reviews, such as title and abstract screening, by automating the identification of relevant studies with high sensitivity. However, their application to full-text screening—a more complex and nuanced task—remains relatively underexplored. This stage requires AI systems to parse entire documents, interpret context, and make decisions aligned with intricate eligibility criteria. Many existing AI tools for systematic reviews often rely on supervised machine learning approaches, requiring extensive training datasets to achieve reliable performance. These tools also exhibit limitations in adaptability, frequently struggling to generalize across diverse topics or study designs. Moreover, validation studies evaluating the accuracy and efficiency of AI tools for full-text screening are limited, underscoring the need for robust research in this domain.

We recently reported findings from a validation study for Loon Lens™, an agentic AI tool designed for efficient level 1 (L1) title and abstract (TiAb) screening in the context of systematic reviews.^5^ That study demonstrated high levels of sensitivity and specificity for L1 screening, and with the incorporation of confidence scores, the most challenging citations could be appropriately directed to human reviewers on the research team. The current study was designed to assess Loon Lens Pro™ for L2 full text screening, building upon our prior work. Unlike traditional approaches, this agentic AI tool requires no prior training and instead operates based on detailed user-defined inclusion and exclusion criteria. By assessing the accuracy, efficiency, and reliability of Loon Lens Pro™ against human reviewers, this research seeks to elucidate the potential of AI to transform systematic review workflows while identifying areas for improvement and further development.

## Methods

### Study Design

This validation study utilized a comparative framework to evaluate the performance of Loon Lens Pro™, an AI-driven full-text screening tool, against human reviewers. The study compared the tool’s decisions with those made by human reviewers, who served as the gold standard, across a sample of full-text articles selected from systematic reviews in diverse fields that formed the basis for our prior validation study for L1 screening.

### AI Tool Description

Loon Lens Pro™ is an agentic AI tool designed to require no training prior to engagement in full text screening. The tool contains multiple AI agents, each specialized in a certain aspect of the screening process. The agents designed within the software’s workflow are inspired by the traditional human-based screening process in systematic reviews, where human reviewers review each article, and document their decisions and thought process. Where disagreements occur, the reviewer agents debate their decisions, and where no consensus is reached, an additional reviewer agent is brought into the process to reach consensus. The tool was designed with outputs that include a clear statement of the rationale that underlies all decisions about article inclusion/exclusion, thereby providing transparency into its screening process. Access to loon Lens Pro™ is publicly available and can be requested on https://loonlens.com/.

### Data Sources and Gold Standard Creation

We previously reported a validation study of our L1 screening tool for titles and abstracts, Loon Lens™, that assessed the tool’s performance against human reviewers across a total of 8 systematic reviews performed by Canada’s Drug Agency (CDA) that were replicated by members of the study team.^6–13^ The studies identified by the search strategies from that work and which were identified as potentially relevant for inclusion were carried forward for full text screening, both using Loon Lens Pro™ and by the study authors (with the latter to serve as the ground truth for this study). Following the removal of conference abstracts as well as citations where corresponding full text articles were behind a paywall, the full texts of the remaining articles were screened. To establish the ground truth decision for all articles, two independent reviewers (GJ, MR) reviewed against the same set of established eligibility criteria as was included in the L1 screening validation study (details are provided in **Supplement 1** and **Supplement 2**). Disagreements were handled through discussion and, if consensus was not reached, a third independent reviewer (BH) was consulted. Final human decision was treated as ground truth. Each AI agent assessed every record based on the full-text, eligibility criteria, and, if applicable, input from other AI agents. A validation step ensured that each record contained the necessary fields for downstream processing.

### Validation Exercise, Data Analysis and Metrics

After screening of all full text articles from the eight chosen SLRs was completed to prepare the set of ground truth decisions of inclusion for each review, full text screening for all articles was next replicated using Loon Lens Pro™. For the full texts associated with each SLR, this process required the submission of the same set of PICOS study selection criteria to Loon Lens Pro™, with full texts downloaded as PDF files and subsequently converted into text files. No training of any form was provided to Loon Lens Pro™ prior to the full text screening task. For all articles associated with each SLR, Loon Lens Pro™ generated a decision of ‘Include’ or ‘Exclude’ (used for validation), along with a corresponding confidence rating for the decision (either ‘Low’, ‘Medium’, or ‘High’; used for the calibration study described below).

We determined the confusion matrix for the screening exercise as a primary means of presenting the screening performance of Loon Lens Pro™. We also estimated the following metrics to assess the performance of Loon Lens Pro™: accuracy (defined as the proportion of correct AI predictions versus ground truth), recall/sensitivity (defined as # true positives [TP]/(# TP + # false negatives), precision (defined as # TP/(#TP + #FP), F1 Score (defined as the harmonic mean of recall and precision), specificity (defined as # TN/(# TN + # false positives [fp]), negative predictive value (NPV; defined as # TN / (# TN + # FN). Bootstrap resampling (1,000 iterations) was used to derive 95% confidence intervals for each metric. On each iteration, we sampled (with replacement) from the results. Where a resampled set lacked both classes, we skipped certain metrics to avoid undefined computations. The 2.5th and 97.5th percentiles of the bootstrap metric distribution were reported as the lower and upper bounds, respectively.

### Confidence Score Assessment

For every screening decision made, Loon Lens Pro™ was designed to provide end users with a confidence score for the decision, labeled as either Low, Medium or High confidence. The functions responsible for this task are based on multiple parameters, including the number of agents used, the sequencing of the agents, and a simulation of multiple runs to estimate the diagnostic results of each unique combination of agents. In the current study, a confidence category for every decision about inclusion/exclusion was captured.

For each full text decision, a binary variable was derived that reflected whether the inclusion decision from Loon Lens Pro™ disagreed with the decision by the team of human reviewers (i.e., the ground truth). Next the confidence category from Loon Lens Pro™ for the decision (i.e., low, medium or high) was coded as a function of two dummy variables (e.g., *confidence_category_low*, *confidence_category_medium*), with the high confidence level serving as the reference category. A logistic regression model was fit to predict the binary outcome error (0 denoted that Loon Lens Pro™ and the human ground truth were aligned, while a value of 1 denoted that they disagreed) using dummy-coded confidence level indicators as predictor variables. Prior to fitting the model, each predictor variable was standardized (mean zero, unit variance) using a StandardScaler from the scikit-learn Python library. The logistic regression was fit on the entire dataset to produce an “apparent” in-sample estimate of performance.

To estimate optimism in the model’s apparent performance, we performed a Harrell’s optimism-correction procedure with 1,000 bootstrap samples. For each bootstrap iteration, a new dataset was constructed by re-sampling (with replacement) the original dataset. Next, we fit a new StandardScaler within the bootstrap sample to avoid data leakage, and subsequently trained a logistic regression model on the scaled bootstrap data. The model’s performance was evaluated on both (a) the bootstrap sample (“in-bag” performance), and (b) the full original dataset. The area under the receiver operating characteristic curve (AUC) was captured on both sets. The difference between the two AUCs (original minus in-bag) yielded an “optimism” estimate for that iteration. This process was repeated 1,000 times, and values were then averaged to estimate the optimism across all bootstraps. The corrected AUC was obtained by subtracting the mean optimism from the apparent AUC computed on the full dataset. Additionally, 95% confidence intervals (CI) were derived for both the apparent AUC and the in-bag-corrected AUC by taking the 2.5th and 97.5th percentiles of the respective bootstrap distributions. We assessed model calibration in-sample by plotting predicted probabilities against observed proportions of error in 20 equally sized probability bins. To further illustrate model output, we generated a histogram of predicted probabilities across the entire dataset.

### Software

All analyses were conducted in Python using the pandas, numpy, scikit-learn, and matplotlib libraries.

## RESULTS

### Overview of Validation Data Source

The eight SLRs that informed both the previously reported L1 TiAb screening validation study as well as the current L2 full text validation study were related to the evaluation of interventions for a range of diagnoses. These conditions consisted of metastatic castration-sensitive prostate cancer^6^, locally advanced or metastatic biliary tract cancer^8^, acute lymphoblastic leukemia and lymphoblastic lymphoma with hypersensitivity to E. coli–derived asparaginase^7^, moderately to severely active ulcerative colitis^9^, psoriatic arthritis^10^, primary hyperoxaluria type 1^11^, severe chronic rhinosinusitis with nasal polyps^12^, and chronic kidney disease with type 2 diabetes.^13^

A total of 3,796 TiAbs were identified by literature searching across the eight SLRs for L1 review, and the team of human reviewers identified a total of 287 (7.6%) citations across reviews to be considered for full text screening. Amongst these, 203 were excluded prior to the L2 validation of Loon Lens Pro™ given they were either conference abstracts or were unavailable as open access articles. This left a total of 84 full text articles for the validation exercise.

### Findings from Validation Exercise

Loon Lens Pro™ showed strong performance in the validation exercise; the confusion matrix summarizing decisions across all full texts is reported in Table 1.

**Table 1:** Table 1: Confusion Matrix, Full Text Screening (n=84)

|  | Predicted Include | Predicted Exclude |
| --- | --- | --- |
| True Include | 18 (21.4%) | 1 (1.2%) |
| True Exclude | 13 (15.5%) | 52 (61.9%) |

Amongst the 84 full texts, totals of 19 (22.6%) and 65 (77.4%) were classified by human reviewers as includes and excludes, respectively. Loon Lens Pro™ correctly classified 70 of the full texts reviewed; 13 (15.5%) were incorrectly predicted as includes, while 1 (1.2%) was incorrectly predicted as an exclude. These predictions produced an accuracy measure of 83.3% (95% CI 75.0%-90.5%), suggesting that the program correctly predicted inclusion at a high rate. The corresponding measures of sensitivity/recall and specificity were 94.7% (95% CI 82.4%-100%) and 80.0% (95% CI 70.1%-89.2%), indicating that Loon Lens Pro™ effectively identified the vast majority of relevant studies (high recall) while also achieving strong specificity, meaning it correctly excluded most irrelevant studies. Loon Lens Pro™ also reached a negative predictive value of 98.1% (95% CI 93.8%%-100%) and a precision of 58.1% (95% CI 41.4%-76.0%). An F1 score of 0.72 (95% CI 0.56-0.84) was also reached, demonstrating the strong performance of Loon Lens Pro for full text screening. All performance metrics are summarized in Figure 1.

### Inspection of AI Rationale for Discrepant Decisions

As described above, totals of 13 false positives and 1 false negative were observed. Rationale provided by Loon Lens Pro™ for its decisions in these cases were reviewed by the study team to establish potential themes in these discrepancies (e.g., ambiguous language, incorrect rationale for decision). In reviewing the provided sets of rationale, trends in the key themes and concepts observed included clear distinction between primary trial publications and secondary analyses, strict adherence to study design criteria, and consideration of how eligibility criteria would need to change to warrant inclusion for borderline cases. Based upon review by study team members, overall, the rationale and insights generated for the full set of 84 full text articles were judged to provide clear insights and interpretations for human members of the review team who may be further appraising full text articles; a sample of rationale generated by Loon Lens Pro™ is provided in **Supplement 3** for interested readers.

**Figure 1:**
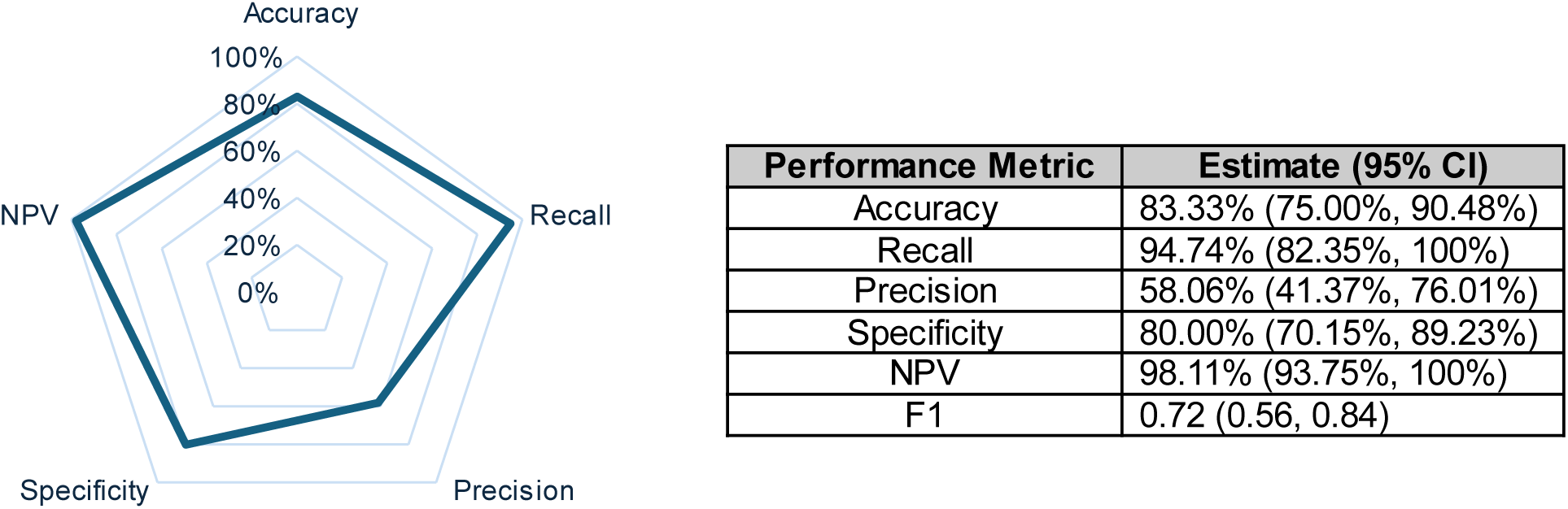
Summary of Performance Metrics NPV = negative predictive value.

### Calibration of Confidence Ratings

“Low” confidence full text articles as determined by Loon Lens Pro ™ (n=16; 19.1% of the sample) were the highest contributors to the overall number of observed errors (n=8; 57.1% of all errors). “Medium” confidence (n=15; 17.9% of the sample) and “high” confidence (n=53; 63.1% of the sample) full texts were responsible for totals of 5 (35.7% of all errors) and 1 (7.1% of all errors) errors, respectively.

The logistic regression analysis performed to approximate the risk of an erroneous decision during full text screening demonstrated the presence of a strong association between the assigned confidence category for each class of citation and the error rates observed. Articles that were categorized as being of a low confidence rating by Loon Lens Pro™ were associated with a predicted error probability of 46.9% (95% CI 22.9%-71.0%). Articles categorized as being of medium and high confidence ratings were associated with predicted error probabilities of 30.9% (95% CI 9.1%-55.2%) and 3.5% (95% CI 1.5%-7.7%), respectively. The regression coefficients from the model each reached statistical significance (p<0.0001), and collectively these findings establish the presence of a negative association between confidence rating and probability of error (i.e., as confidence of Loon Lens Pro ™ decisions increase, the probability of an error is lowered). A C index of 0.87 (95% CI 0.76, 0.94) was found, demonstrating a strong ability of the model to distinguish between correct and incorrect full text screening decisions by Loon Lens Pro ™. The apparent C-index was 0.8561. Bootstrapping over 1,000 iterations yielded a mean optimism estimate of 0.0152, resulting in a corrected C-index of 0.8409 (95% CI, 0.7184–0.9447).

## DISCUSSION

In this study, we evaluated the performance of Loon Lens Pro™—an agentic AI tool designed for full-text (L2) screening in systematic literature reviews—against decisions made by human reviewers. Our findings demonstrate that Loon Lens Pro™ achieved a high sensitivity (94.7%) and strong overall accuracy (83.3%), while maintaining respectable specificity (80.0%) and negative predictive value (98.1%). These results underscore the tool’s capability to reliably identify relevant studies in the context of full-text screening, a phase in the SLR process known to be particularly resource intensive and time consuming. Prior estimates suggest that screening activities can consume approximately 25% or more of the total effort in systematic reviews, and by automating L2 screening, tools like Loon Lens Pro™ hold great promise for significantly reducing reviewer workload.

The current study highlights the potential of AI tools such as Loon Lens Pro™, an autonomous agentic AI platform, to streamline full-text screening in systematic reviews. Loon Lens Pro™ demonstrated high sensitivity and substantial agreement with human reviewers, underscoring its reliability in replicating human decisions. The involvement of confidence scores to qualify the degree of challenge for each AI decision about inclusion also were found to provide value in considering the allocation of challenging citations to members of the human review team. This approach fits well with the workflow of human/machine review workflows that many have begun to envision for future work. Tools to expedite both TiAb and full text screening through application of AI will almost certainly drive more and more knowledge synthesis initiatives in the future given the ever-increasing pressures for living/’immediately available’ evidence.

To our knowledge, this is the first publicly available automated tool to address the task of full text screening. The automation of full text screening was technically unattainable until the advancements of large language models. Traditionally, the number of full-text articles to screen would not have been sufficient to train a project-specific machine learning model with an Active Learning framework, which represents the main approach in L1 screening where the user has to screen a number of articles in order to train a machine learning model. However, with the more limited number of studies typically included for L2 screening, as compared to the number of citations in L1 screening, active learning approaches would not be feasible in this task. Large language and other foundational models are pre-trained on a large corpus of data that allows emergent properties such as reasoning and flexibility in task applications. Agentic AI approaches, in which multiple large language models—each assigned a specific task, arranged in a structured workflow, and equipped with capabilities such as tool use—have enabled the orchestration of generalized foundational models in a way that simulates human teams, often without additional fine-tuning.

Combined with our previously published validation work of Loon Lens™ for L1 screening^5^, this work represents another important step toward substantially increasing the efficiency of evidence synthesis and systematic reviews, allowing for more feasible living systematic reviews and clinical guidelines. The ability to automate both phases of the study selection process—even partially—can translate into substantial time and resource savings and facilitates improved SLR workflows. By flagging low-confidence decisions for human review, the tool not only accelerates the screening process, but also helps maintain the rigor and reproducibility of systematic reviews. As the volume of scientific literature continues to grow, such AI-driven approaches will become increasingly critical in ensuring that evidence synthesis keeps pace with the expanding knowledge base.

Loon Lens Pro™ represents a scientifically validated approach to incorporate generative AI into the systematic review process. Loon Lens Pro™ is a specialized tool that removes several challenges from researchers regarding building, developing, and validating tools for individual systematic review projects. Instead, this tool can be incorporated in various ways within the traditional SLR process. For example, Loon Lens Pro™ can act as the second independent reviewer in the screening process, saving substantial resources on each project. Loon Lens Pro™ can also be used by incorporating it into a human-in-the-loop process where a human reviewer would review certain number of citations or specific categories of the output. For example, a human reviewer may decide to only review output that is determined to have Low or Medium confidence levels, or a randomly selected sample of the output to verify the quality and accuracy of decisions.

A key strength of Loon Lens Pro™ lies in its high recall. By correctly identifying the vast majority of relevant articles, the tool minimizes the risk of excluding studies that could ultimately influence the conclusions of a review. While the precision was observed to be moderate (58.1%), this is an acceptable trade-off in a screening context where the primary aim is to maximize sensitivity. In practice, the presence of false positives—approximately 15.5% of full texts in our sample—can be managed through human-in-the-loop verification. Notably, the AI-generated descriptions of judgement rationale provide clear insights into its decision-making process, mimicking the nuanced considerations of human reviewers. The descriptions of rationale not only enhance trust in the automated process, but also help reviewers understand borderline cases and discrepancies in decision-making.

One of the most compelling aspects of Loon Lens Pro™ is its built-in confidence scoring mechanism. The logistic regression analysis revealed a strong negative association between the tool’s confidence ratings and the probability of error: full texts with a low confidence rating were associated with nearly a 47% predicted error probability, whereas those with a high confidence rating exhibited an error probability of only 3.5%. The regression model demonstrated robust discriminative ability (C-index of 0.87, with a corrected C-index of 0.84 after minimal optimism correction). This calibrated confidence measure serves as an important decision support feature, allowing human reviewers to prioritize articles flagged as low confidence for additional scrutiny. In doing so, the tool not only expedites the screening process but also enhances overall review quality by directing expert attention where it is most needed.

### Limitations, Strengths and Future Directions

There are limitations of this work that are worthy of mention. First, the sample of full text articles used for this validation study were derived from a set of eight systematic reviews and encompassed a total of only 84 articles. While the articles were diverse in topics and were selected using a systematic approach, they may not fully represent the complexity of SLRs generally and are of limited sample size. In the future we intend to conduct a larger validation study that will encompass additional systematic reviews of diverse topics and we will expand the types of eligible studies from purely RCTs to others such as observational studies and health-economic studies, thereby seeking to broaden the generalizability of Loon Lens Pro™. These will broaden our understanding of the performance of Loon Lens Pro™ for more diverse tasks of study selection.

There are also important strengths that should be noted. First, this work demonstrated the potential of Loon Lens Pro™ to operate efficiency for full text screening. This represents an important solution for systematic reviews that can help reduce costs and improve efficiencies which has long been sought after and which leverages the strengths of available LLM technologies; this can have important implications for various applications including the performance of de novo reviews, systematic review updates, living systematic reviews and other such applications. The transparency of Loon Lens Pro™ in providing rationale for its decisions and in categorizing the related level of confidence also feeds well into the collaborative nature of “human in the loop” reviews, an approach which will be pivotal to working with AI technologies for SLR performance in the immediate future.^14,15^ Next, we employed a robust statistical approach involving logistic regression and optimism correction that demonstrated excellent calibration and performance to predict the risk of incorrect full text screening errors; the incorporation of Loon Lens Pro™ into systematic review workflows is highly feasible.

## Conclusions

Our study provides evidence that Loon Lens Pro™ is a viable and effective tool for automating the full-text screening phase of systematic reviews. Its high sensitivity, robust confidence scoring mechanism, and transparent rationale generation collectively support its potential to alleviate the burden of manual screening without compromising the quality of study selection.

## Data Availability

Access to raw data that informed this study are available upon request from the authors.

## Article Information

### Author contributions

GJ and MU conceptualized the study. GJ, MU, MJ designed and developed Loon Lens ProTM. GJ and MU conducted all screening involving Loon Lens ProTM. GJ was responsible for data analyses. GJ, MU, and BH substantially contributed to the interpretation of the results. GJ and BH prepared the preliminary draft of the manuscript. All co-authors reviewed the final version of the manuscript and provided final approval.

### Ethics

No ethics approval was required for the current study.

### Data Availability

All results are provided within the manuscript. Access to raw data that informed this study are available upon request from the authors.

## Appendices

**Appendix 1:** Literature Searches for Systematic Reviews

**Appendix 2:** SLR Eligibility Criteria

**Appendix 3:** Sample Rationale Produced by Loon Lens Pro™

## Appendix 1

### Literature Searches for Chosen Systematic Reviews

#### PC0294 Nubeqa

((darolutamide OR “darolutamide”) OR (nubeqa OR “nubeqa”) OR (“darramamide” OR darramamide) OR (“bay-1841788” OR “bay1841788” OR “odm-201” OR “odm201” OR “orm-16497” OR “orm16497” OR “orm-16555” OR “orm16555” OR “X05U0N2RCO”)) AND (((randomized OR randomised) AND (trial OR study OR research OR clinical OR controlled)) OR (phase AND (I OR II OR III OR IV OR 1 OR 2 OR 3 OR 4 OR one OR two OR three OR four)) OR “placebo” OR “double-blind” OR “double_blind” OR “double blind” OR “open label” OR “open-label” OR “open_label” OR “single-blind” OR “single_blind” OR “single blind”)

#### PC0296 Imfinzi

(((((bile OR hepatic OR gall OR gallbladder OR (gall AND bladder)) AND (duct OR tract)) OR hepatobiliary OR hepatocellular OR choledochus OR colangio OR ampulla) AND (cancer OR tumor OR tumour OR carcinoma OR neoplasm OR malignant OR adenocarcinoma)) OR cholangiocarcinoma) AND ((durvalumab OR “durvalumab”) OR (imfinzi OR “imfinzi”) OR (“medi 4736” OR “medi4736”)) AND (((randomized OR randomised) AND (trial OR study OR research OR clinical OR controlled)) OR (phase AND (I OR II OR III OR IV OR 1 OR 2 OR 3 OR 4 OR one OR two OR three OR four)) OR “placebo” OR “double-blind” OR “double_blind” OR “double blind” OR “open label” OR “open-label” OR “open_label” OR “single-blind” OR “single_blind” OR “single blind”)

#### PC0301 Rylaze

((“Asparaginase” OR Asparaginase) OR (“rylaze” OR rylaze) OR (“crisantaspas” OR crisantaspas) OR (“erwinase” OR erwinase) OR (“erwinaze” OR erwinaze) OR (“jzp 458” OR “jzp458” OR “D733ET3F9O”)) AND (((randomized OR randomised) AND (trial OR study OR research OR clinical OR controlled)) OR (phase AND (I OR II OR III OR IV OR 1 OR 2 OR 3 OR 4 OR one OR two OR three OR four)) OR “placebo” OR “double-blind” OR “double_blind” OR “double blind” OR “open label” OR “open-label” OR “open_label” OR “single-blind” OR “single_blind” OR “single blind”)

#### SR0730 Rinvoq

((ulcerative AND colitis) OR (“ulcerative colitis”) OR (Colitis OR “colitis”) OR (proctocolitis OR “proctocolitis”) OR (colorectitis OR “colorectitis”)) AND ((“rinvoq” OR rinvoq) OR (“upadacitinib” OR upadacitinib) OR (“abt494” OR “abt 494” OR “NEW4DV02U5”))

#### SR0733 Tremfya

((tremfya OR “tremfya”)OR (guselkumab OR “guselkumab”) OR (“cnto 1959” OR “cnto1959” OR “089658A12D”)) AND (((randomized OR randomised) AND (trial OR study OR research OR clinical OR controlled)) OR (phase AND (I OR II OR III OR IV OR 1 OR 2 OR 3 OR 4 OR one OR two OR three OR four)) OR “placebo” OR “double-blind” OR “double_blind” OR “double blind” OR “open label” OR “open-label” OR “open_label” OR “single-blind” OR “single_blind” OR “single blind”)

#### SR0734 Oxlumo

((lumasiran OR “lumasiran”) OR (Oxlumo OR “Oxlumo”) OR (“ad 65585” OR “ad65585” OR “aln 65585” OR “aln65585” OR “aln g01” OR “alng01” OR “aln go1” OR “alngo1” OR “RZT8C352O1” OR “67P6XH37HD”))

#### SR0735 Nucala

((((rhino AND sinusitis) OR (rhinosinusitis) OR (sinus AND inflammation) OR Sinusitis OR rhinitis) AND (chronic OR persistent OR recurrent OR flareup OR “flare up”)) OR ((nasal AND polyp) OR rhinopolyp OR “CRSwNP”)) AND ((Nucala OR “Nucala”) OR (mepolizumab OR “mepolizumab”) OR (bosatria OR “bosatria”) OR (“90Z2UF0E52” OR “SB240563” OR “SB 240563”)) AND (((randomized OR randomised) AND (trial OR study OR research OR clinical OR controlled)) OR (phase AND (I OR II OR III OR IV OR 1 OR 2 OR 3 OR 4 OR one OR two OR three OR four)) OR “placebo” OR “double-blind” OR “double_blind” OR “double blind” OR “open label” OR “open-label” OR “open_label” OR “single-blind” OR “single_blind” OR “single blind”)

#### SR0737-Kerendia

((finerenone OR “finerenone”) OR (kerendia OR “kerendia”) OR (“BAY94-8862” OR “BAY-94-8862” OR “BAY948862” OR “BAY-948862” OR “DE2O63YV8R”)) AND (((randomized OR randomised) AND (trial OR study OR research OR clinical OR controlled)) OR (phase AND (I OR II OR III OR IV OR 1 OR 2 OR 3 OR 4 OR one OR two OR three OR four)) OR “placebo” OR “double-blind” OR “double_blind” OR “double blind” OR “open label” OR “open-label” OR “open_label” OR “single-blind” OR “single_blind” OR “single blind”)

## Appendix 2

### SLR Eligibility Criteria

#### PC0294 Nubeqa

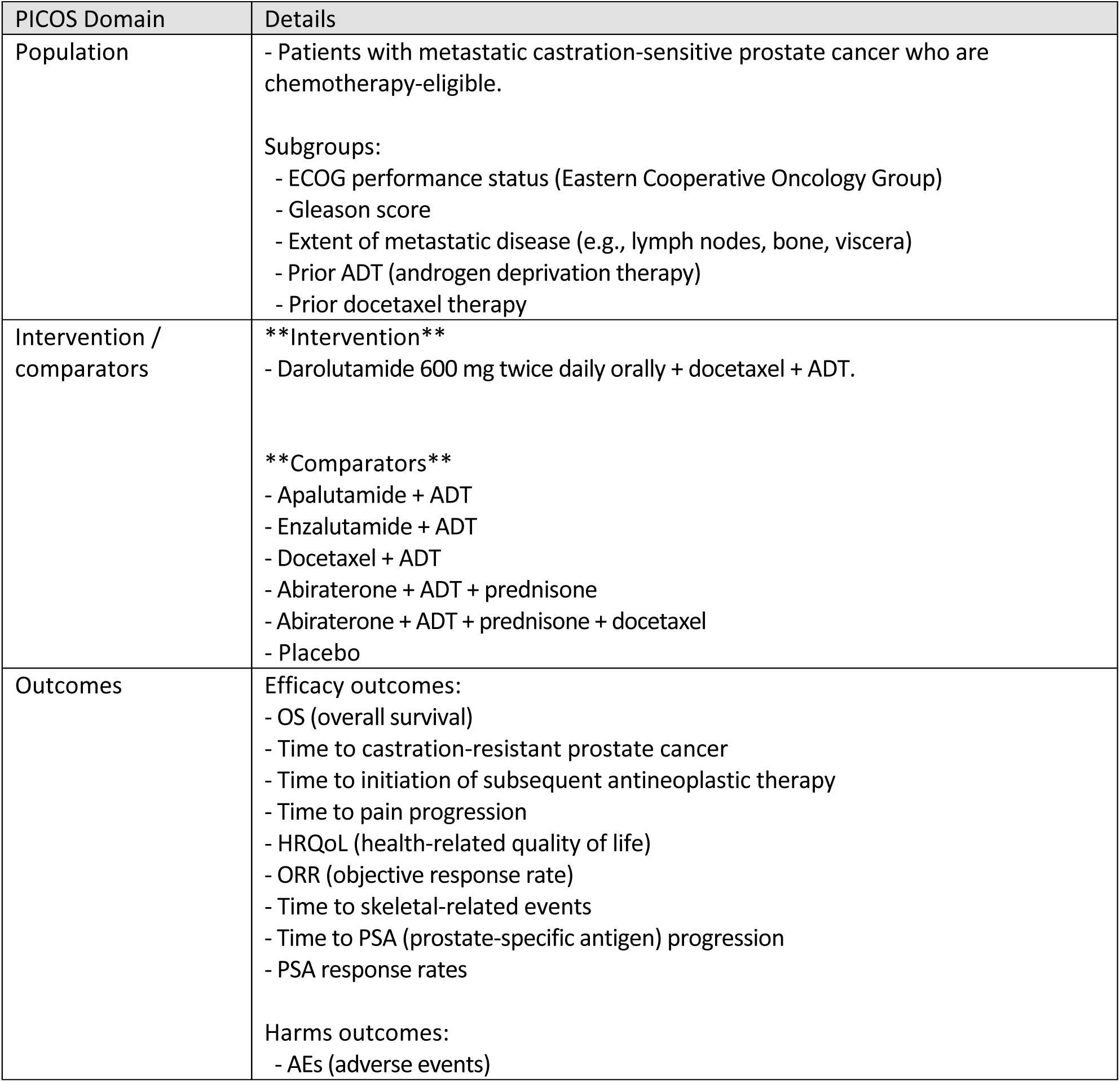
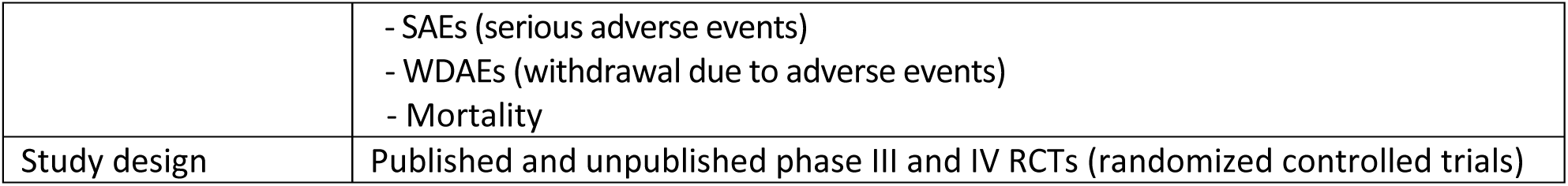

#### PC0296 Imfinzi

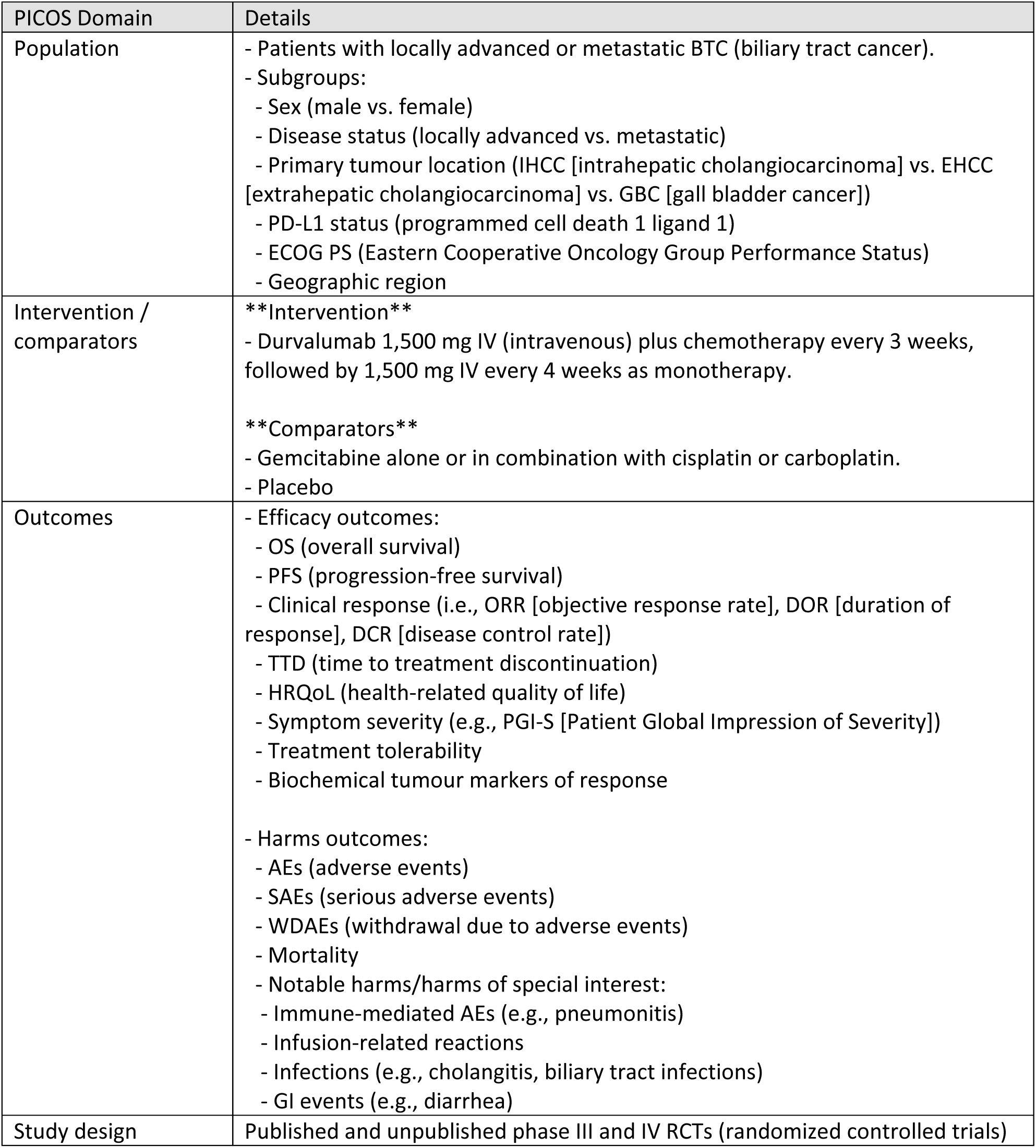

#### SR0730 Rylaze

PC0301 Rylaze
| PICOS Domain | Details |
| --- | --- |
| Population | - Adult and pediatric populations aged 1 year and older receiving a multidrug chemotherapeutic regimen for the treatment of ALL (acute lymphoblastic leukemia) and LBL (lymphoblastic lymphoma) who have developed hypersensitivity to E. coli–derived asparaginase.<br>- Subgroups:<br>- Age group (e.g., < 25 years vs. ≥ 25 years)<br>- ALL vs. LBL<br>- CNS (central nervous system) involvement |
| Intervention / comparators | **\*\*Intervention\*\***<br>- Crisantaspase recombinant 25 mg/m<sup>2</sup> on Monday and Wednesday and 50 mg/m<sup>2</sup> on Friday, administered intramuscularly, for a total of 6 doses (to replace each planned dose of long-acting E. coli–derived asparaginase).<br>**\*\*Comparators\*\***<br>- Erwinia-derived asparaginase<br>- No comparator |
| Outcomes | - Efficacy outcomes:<br>- Overall survival<br>- Event-free survival<br>- Disease-free survival<br>- Complete clinical remission and/or minimal residual disease<br>- NSAA (nadir serum asparaginase activity) levels<br>- HRQoL (health-related quality of life)<br>- Harms outcomes:<br>- AEs (adverse events), SAEs (serious adverse events), and WDAEs (withdrawal due to adverse events)<br>- Mortality<br>- Notable harms: thrombosis, pancreatitis, hemorrhage, hypersensitivity reaction, and hepatotoxicity |
| Study design | - Phase II and Phase III RCTs (randomized controlled trials) and single arm trials |

#### SR0730 Rinvoq

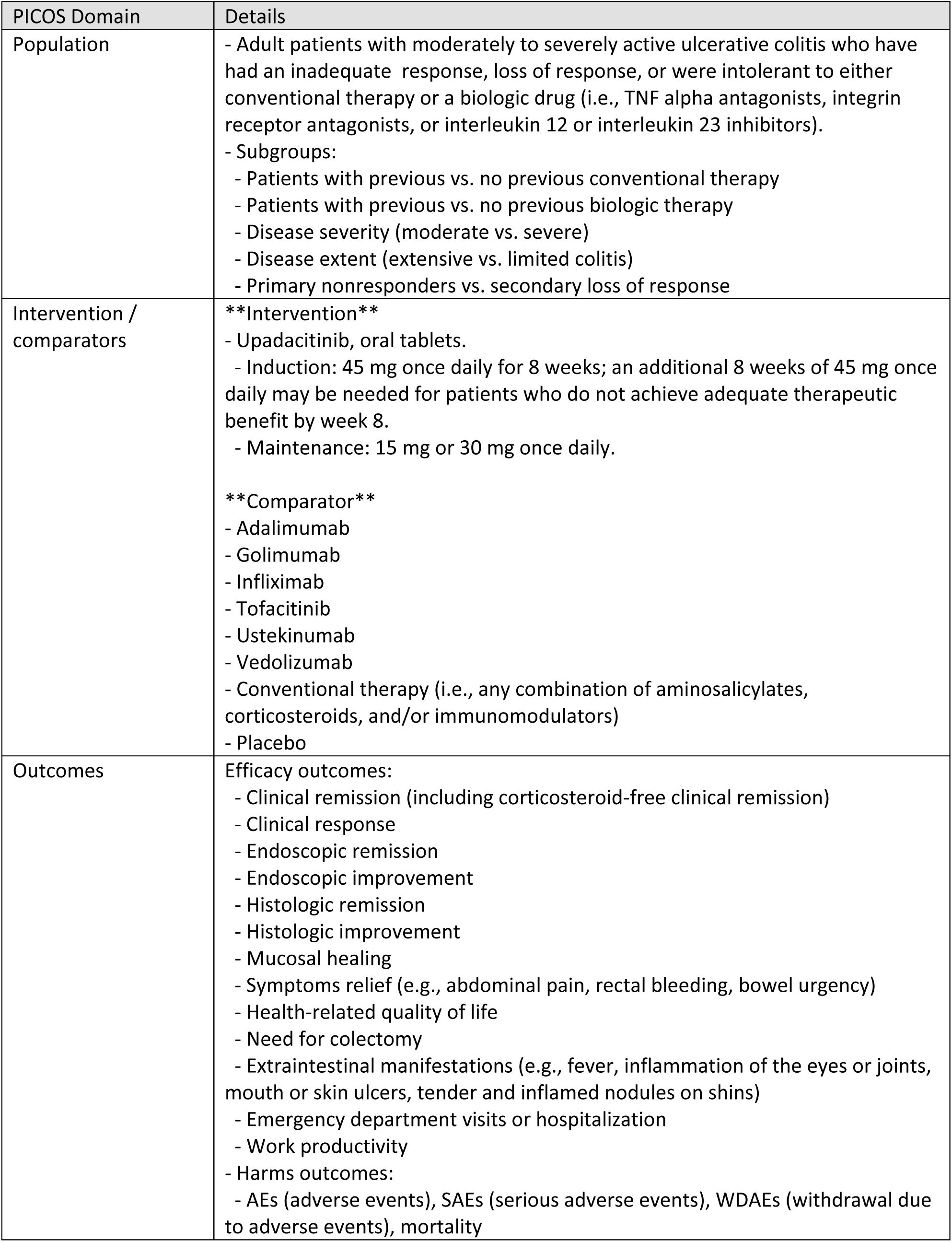
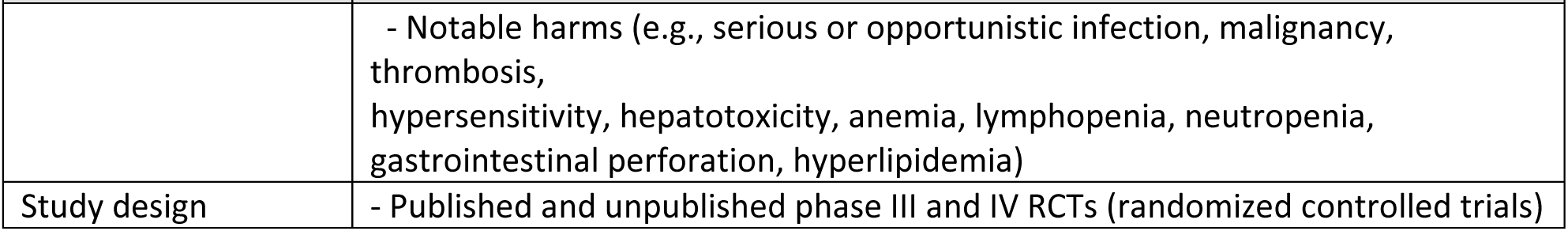

| PICOS Domain | Details |
| --- | --- |
| Population | - Adult patients with moderately to severely active ulcerative colitis who have had an inadequate response, loss of response, or were intolerant to either conventional therapy or a biologic drug (i.e., TNF alpha antagonists, integrin receptor antagonists, or interleukin 12 or interleukin 23 inhibitors).<br>- Subgroups:<br>- Patients with previous vs. no previous conventional therapy<br>- Patients with previous vs. no previous biologic therapy<br>- Disease severity (moderate vs. severe)<br>- Disease extent (extensive vs. limited colitis)<br>- Primary nonresponders vs. secondary loss of response |
| Intervention / comparators | **\*\*Intervention\*\***<br>- Upadacitinib, oral tablets.<br>- Induction: 45 mg once daily for 8 weeks; an additional 8 weeks of 45 mg once daily may be needed for patients who do not achieve adequate therapeutic benefit by week 8.<br>- Maintenance: 15 mg or 30 mg once daily.<br>**\*\*Comparator\*\***<br>- Adalimumab<br>- Golimumab<br>- Infliximab<br>- Tofacitinib<br>- Ustekinumab<br>- Vedolizumab<br>- Conventional therapy (i.e., any combination of aminosalicylates, corticosteroids, and/or immunomodulators)<br>- Placebo |
| Outcomes | Efficacy outcomes:<br>- Clinical remission (including corticosteroid-free clinical remission)<br>- Clinical response<br>- Endoscopic remission<br>- Endoscopic improvement<br>- Histologic remission<br>- Histologic improvement<br>- Mucosal healing<br>- Symptoms relief (e.g., abdominal pain, rectal bleeding, bowel urgency)<br>- Health-related quality of life<br>- Need for colectomy<br>- Extraintestinal manifestations (e.g., fever, inflammation of the eyes or joints, mouth or skin ulcers, tender and inflamed nodules on shins)<br>- Emergency department visits or hospitalization<br>- Work productivity<br>Harms outcomes:<br>- AEs (adverse events), SAEs (serious adverse events), WDAEs (withdrawal due to adverse events), mortality |
|  | - Notable harms (e.g., serious or opportunistic infection, malignancy, thrombosis, hypersensitivity, hepatotoxicity, anemia, lymphopenia, neutropenia, gastrointestinal perforation, hyperlipidemia) |
| Study design | - Published and unpublished phase III and IV RCTs (randomized controlled trials) |

#### SR0733 Tremfya

| PICOS Domain | Details |
| --- | --- |
| Population | - Adults with active psoriatic arthritis.<br>- Subgroups:<br>- Previous exposure to bDMARDs (biologic disease-modifying antirheumatic drugs) (e.g., treatment-naïve; treatment-experienced; non-responsive or intolerant)<br>- Concomitant treatment with non-biologic DMARDs |
| Intervention / comparators | **\*\*Intervention\*\***<br>- Guselkumab 100 mg SC (subcutaneous) injection at week 0 and week 4, followed by maintenance dosing every 8 weeks thereafter (alone or in combination with a cDMARD [e.g., methotrexate]).<br>**\*\*Comparators\*\***<br>- TNF inhibitors (adalimumab, certolizumab pegol, etanercept, golimumab, infliximab)<br>- IL-17 inhibitors (ixekizumab, secukinumab)<br>- IL-12/23 inhibitors (ustekinumab)<br>- IL-23 inhibitors (risankizumab)<br>- JAK inhibitors (tofacitinib, upadacitinib)<br>- Apremilast<br>- Abatacept<br>- cDMARDs (e.g., methotrexate) alone or in combination with biologic or other DMARDs<br>- Placebo |
| Outcomes | - Efficacy outcomes:<br>- Clinical response in PsA (psoriatic arthritis) symptoms (e.g., ACR 20, ACR 50, ACR 70, MDA, DAS 28)<br>- Measure of function and disability (e.g., HAQ-DI)<br>- Health-related quality of life<br>- Measure of skin disease (e.g., PASI 75, PASI 90, PASI 100, or IGA response)<br>- Measure of other musculoskeletal disease (e.g., dactylitis, enthesitis, and axial arthritis)<br>- Measure of PsA symptoms (e.g., pain, fatigue)<br>- Radiologic changes<br>- Harms outcomes:<br>- AEs (adverse events), SAEs (serious adverse events), WDAEs (withdrawal due to adverse events), mortality, and notable harms (serious infections, hypersensitivity reactions, elevated hepatic enzymes, hepatic disorders, injection-site reactions) |
| Study design | - Published and unpublished phase III and IV RCTs (randomized controlled trials) |

#### SR0734 Oxlumo

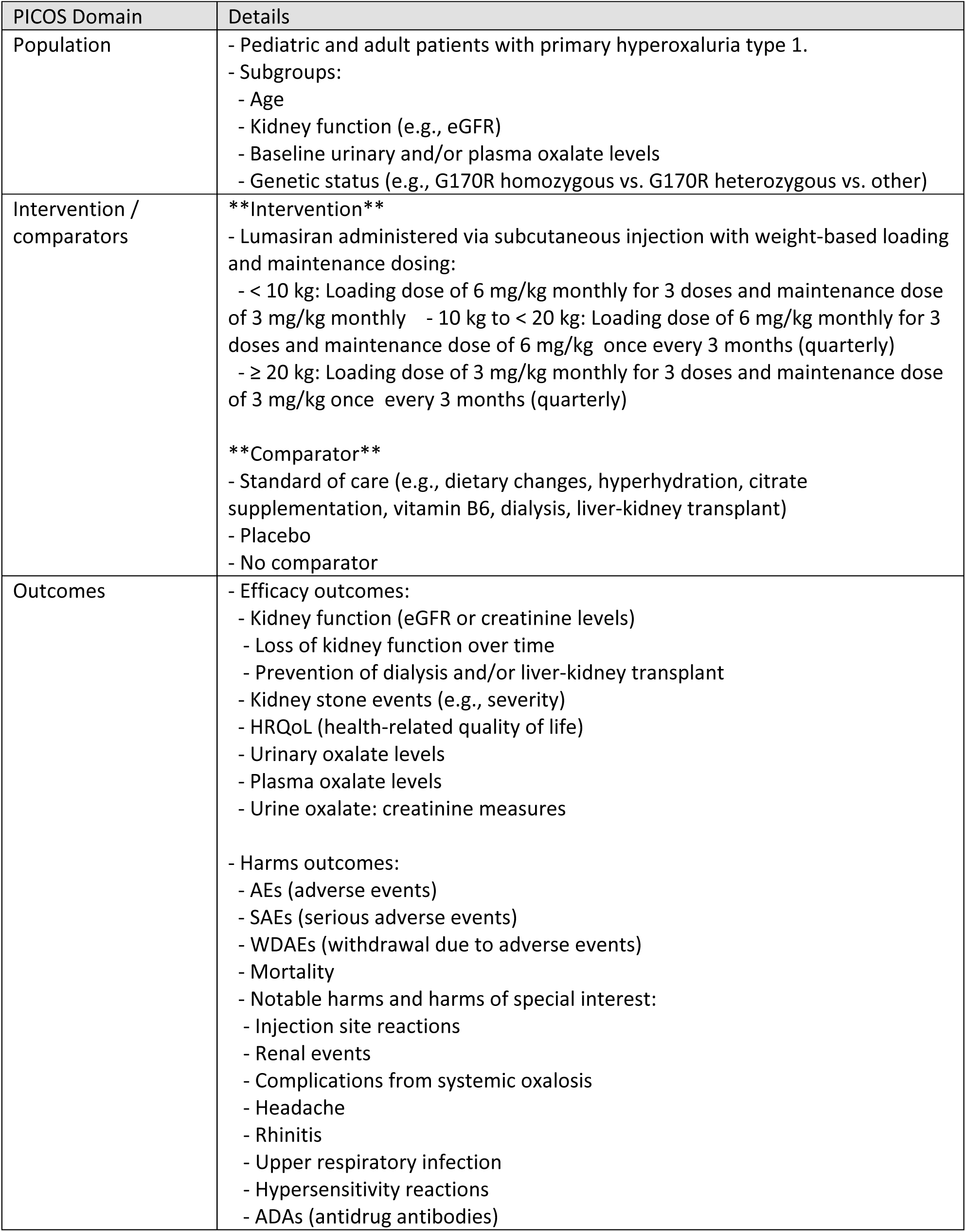
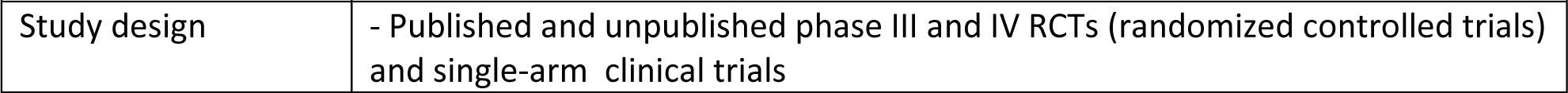

#### SR0735 Nucala

| PICOS Domain | Details |
| --- | --- |
| Population | - Adult patients 18 years and older with severe CRSwNP (chronic rhinosinusitis with nasal polyps) inadequately controlled by intranasal corticosteroids alone.<br>- Subgroups:<br>- Asthma diagnosis (yes/no)<br>- Prior surgery (yes/no) |
| Intervention / comparators | **\*\*Intervention\*\***<br>- Mepolizumab, 100 mg administered by SC (subcutaneous) injection once every 4 weeks, used in combination with intranasal corticosteroids and/or saline irrigation.<br>**\*\*Comparator\*\***<br>- Intranasal corticosteroids and/or saline irrigation.<br>- Placebo |
| Outcomes | **\*\*Outcomes\*\***<br>- Efficacy outcomes:<br>- Nasal obstruction<br>- VAS (visual analogue scale) for nasal obstruction<br>- Symptoms:<br>- Composite VAS symptom score for nasal discharge, feeling of mucus in the throat, loss of smell, facial pain, and nasal polyp symptoms<br>- Sense of smell<br>- Response to treatment:<br>- Change in nasal polyp size<br>- Severity of nasal polyps and nasal obstruction:<br>- Endoscopic nasal polyp score<br>- Nasal congestion:<br>- PnIF (peak nasal inspiratory flow)<br>- HRQoL (health-related quality of life):<br>- SNOT-22 (Sino-Nasal Outcome Test 22)<br>- Systemic steroid use for nasal polyps<br>- Nasal inflammation:<br>- CT imaging<br>- Nasal polyp surgery:<br>- Need for nasal surgery<br>- Time to first nasal surgery<br>- Nasal surgery at 24 weeks<br>- Work productivity:<br>- WPAI-GH (Work Productivity and Activity Impairment — General Health)<br>- Harms outcomes:<br>- AEs (adverse events)<br>- SAEs (serious adverse events)<br>- WDAEs (withdrawal due to adverse events)<br>- Mortality |
|  | - Notable harms of special interest, including systemic and local injection site reactions; serious and opportunistic infections; serious cardiac, vascular, and thromboembolic events; and serious ischemic events. |
| Study design | - Published and unpublished phase III and IV RCTs (randomized controlled trials) |

#### SR0737-Kerendia

| PICOS Domain | Details |
| --- | --- |
| Population | - Adults with chronic kidney disease and type 2 diabetes.<br>- Subgroups:<br>- Albuminuria at baseline<br>- eGFR (estimated glomerular filtration rate) at baseline<br>- SGLT2 (sodium-glucose cotransporter-2) inhibitor use at baseline<br>- History of cardiovascular disease<br>- Use of ACE (angiotensin-converting enzyme) inhibitor and/or ARB (angiotensin receptor blocker) |
| Intervention / comparators | **\*\*Intervention\*\***<br>- Finerenone 10 mg and 20 mg, oral administration.<br>**\*\*Comparators\*\***<br>- Placebo plus SOC (standard of care, including an ACE inhibitor or ARB)<br>- SGLT2 inhibitor plus SOC |
| Outcomes | **\*\*Outcomes\*\***<br>- Efficacy outcomes:<br>- Renal events (e.g., kidney failure)<br>- eGFR (estimated glomerular filtration rate)<br>- Urinary albumin-creatinine ratio<br>- Cardiovascular events (e.g., myocardial infarction)<br>- Mortality (renal, cardiovascular, and all-cause)<br>- Hospitalization (renal, cardiovascular, and all-cause)<br>- HRQoL (health-related quality of life)<br>- Symptom severity<br>- Functional status<br>- Harms outcomes:<br>- AEs (adverse events)<br>- SAEs (serious adverse events)<br>- WDAEs (withdrawal due to adverse events)<br>- Mortality<br>- Notable harms or harms of special interest (e.g., hyperkalemia, new onset of atrial fibrillation and atrial flutter, hypotension) |
| Study design | - Published and unpublished phase III and IV RCTs (randomized controlled trials) |

## Appendix 3

### Sample Rationale Produced by Loon Lens Pro™

The reasoning provided by Loon Lens Pro™ provides a description of the platform’s assessment of the citation in question with regard to factors to be considered related to the a priori selection criteria that will influence the final decision about inclusion. This information will be helpful to supporting human reviewers with rationale for the decisions made, as well as for the handling of lower confidence abstracts where human members of the research team perform targeted screening. An example of an evaluation for an abstract is provided below from one of the included reviews of an intervention for metastatic castration-sensitive prostate cancer.

**\*\*Analysis of the study against the inclusion criteria:\*\***

**\*\*Population:\*\*** The study includes patients with metastatic, hormone-sensitive prostate cancer who are eligible for docetaxel and ADT. This matches the inclusion criteria.

**\*\*Intervention:\*\*** The intervention is darolutamide 600 mg twice daily + docetaxel + ADT. This matches the inclusion criteria.

**\*\*Comparator:\*\*** The comparator is placebo + docetaxel + ADT. While placebo is not explicitly listed as a comparator in the inclusion criteria, it represents the control arm for assessing the effect of darolutamide and is therefore relevant.

**\*\*Outcomes:\*\*** The study assesses overall survival as the primary endpoint, which is listed in the inclusion criteria. It also evaluates several secondary endpoints that align with the inclusion criteria (e.g., time to castration-resistant prostate cancer, time to pain progression).

**\*\*Study Design:\*\*** The study is a phase 3 randomized controlled trial (RCT). This matches the inclusion criteria.

**\*\*Arguments for inclusion:\*\***

* The study directly addresses the research question of the systematic review by investigating the effect of darolutamide in combination with docetaxel and ADT in men with metastatic, hormone-sensitive prostate cancer.

* The study design, population, intervention, and primary outcome all align with the predefined inclusion criteria.

**\*\*Arguments for exclusion:\*\***

* None. All aspects of the study appear to meet the inclusion criteria.

**\*\*Conclusion:\*\***

Based on the analysis above, this study should be *included* in the systematic review. It provides high-quality evidence relevant to the research question and meets all predefined inclusion criteria.

